# The implementation of an unscheduled care co-ordination hub (Flow Navigation Centre Plus), and emergency department attendances and delays: a controlled interrupted time series

**DOI:** 10.64898/2026.08.28.26361651

**Authors:** Ryan D McHenry, Christopher EJ Moultrie

## Abstract

**Objectives:** Emergency Department (ED) crowding is an international concern, predominantly caused by ‘exit block’, the lack of availability of inpatient beds for those requiring admission. The implementation of Flow Navigation Centre Plus (FNC+) services in Scotland aimed to reduce self-presentation to EDs and reduce crowding by providing remote clinical assessment for patients contacting urgent care by telephone and professional-to-professional advice on patient pathways, but their effectiveness is unknown. This study aimed to estimate the effect of board-wide implementation of FNC+ on ED attendances and long waits during the first year of FNC+ operation.

**Methods:** Controlled interrupted time series using weekly, publicly reported Public Health Scotland data. The intervention was implementation of the FNC+ in NHS Lanarkshire on 1 April 2024. Counts were summed across constituent sites and percentages derived from board totals. Co-primary outcomes were ED attendance volume and the proportions of attendances spending more than 4, 8 and 12 hours in the department. Segmented regression was fitted with contemporaneous control boards, seasonal terms, and accounted for autoregression.

**Results:** 118 pre-intervention and 52 post-intervention weeks were analysed across all 3 EDs in the implementing board. Attendances showed no detectable step change (+1.20%; 95%CIs −0.66 to +3.10) relative to the counterfactual. The estimated effect increased across follow-up, however, changing by +3.95% over 52 weeks (95% CI +0.36 to +7.67%). There was no significant step change in the proportion of attendances waiting more than 4 hours following the intervention (+1.74%; 95%CIs −0.71 to 4.20%). Some transition and structural sensitivity analyses demonstrated significant deteriorations in ED performance, and increased attendances, in the year following implementation, and none demonstrated improvements.

**Conclusions:** Board-wide implementation of a Flow Navigation Centre Plus was not associated with a step change in ED attendances or in long waits, but there is some evidence that attendances increased and long waits increased in the year following implementation. Their provision of supply-sensitive care is a possible mechanism. Additionally, given their action at the point of input, aiming to divert patients from ED attendance, it is unlikely that such services could relieve a constraint due to exit block, the availability of inpatient care for those requiring admission.

## Introduction

Emergency department (ED) crowding is an international public health concern and is associated with patient harm, including excess mortality.[1] Crowding is principally attributed to output failures: the lack of timely access to inpatient beds for patients requiring admission, also termed “exit block”.[2,3]

Exit block is driven by delayed discharges, prolonged length of stay and high occupancy within hospital inpatient wards.[4] Despite this, health systems internationally have attempted to mitigate ED crowding and its harms through interventions designed to reduce demand for emergency care.[4] Such approaches have been undertaken in Scotland during the Redesign of Urgent Care programme, [5] which established Flow Navigation Centres (FNCs) in every regional health board.

FNCs were originally conceived to direct attendances from unscheduled emergency care to local services, and facilitate rapid access to a senior clinical decision maker by telephone (provided through existing NHS 24 telephone health advice pathways) or digitally. This was expected to provide a remote clinical consultation or onward specialist referral without the need for patients to attend the ED. The service has since been expanded in some regions, branded as a Flow Navigation Centre Plus (FNC+)[6] to provide the same core services alongside professional-to-professional advice for primary care and ambulance service clinicians, facilitating access to alternative patient pathways and diverting presentations to unscheduled care.[5]

There is no currently available evidence on the extent to which the introduction of such services affects patient flow in emergency care. Improvements may reasonably be expected where senior clinical assessment at first contact can identify and divert patients who do not require assessment in emergency care; or schedule those who can attend less urgently to mitigate arrival peaks that contribute to crowding.[4,7] However, there is evidence that targeting intervention at a point distinct from the constraint of a system may have little or no effect on outcomes,[7,8] and FNCs/FNC+ may act in a way consistent with supply-sensitive care, where the availability of a service itself contributes to its utilisation.[9] In such a system it might be expected that some contacts will substitute an attendance to emergency care, but that others will be generated by the existence of the service, among people who would otherwise have waited, self-managed or consulted elsewhere, and a proportion of those will be directed onward to the ED. [10] Previous evidence, on the implementation of telephone health advice services in the UK, has shown results consistent with this mechanism.[11]

This study aimed to provide evidence on the effectiveness of FNC+ by estimating the effect of board-wide implementation of an FNC+ on overall patient flow, using ED attendance volume and the proportions of patients waiting more than 4, 8 and 12 hours.

## Methods

### Setting and intervention

This evaluation considers FNC+ as implemented in NHS Lanarkshire, an NHS Scotland territorial health board operating three Type 1 EDs (University Hospital Hairmyres, University Hospital Monklands, University Hospital Wishaw). Type 1 EDs are typically larger departments, providing 24-hour Consultant-led services.[12] With first-year funding of £2,791,268 the core additions to the existing FNC service included:[13]

1. An expansion of referral sources to include General Practitioners (GPs), ambulance clinicians, police, and mental health services, in addition to existing referral sources from NHS 24.
2. Expansion of patient disposition options, including direction of patients to community respiratory services, Hospital at Home, mental health services, pharmacy services and out-of-hours services.
3. An expansion of multidisciplinary staffing including nurses, administrative staff, Emergency Medicine consultants and GPs.
4. System-level demand management.[14,15]

The service was launched at the beginning of April 2024 as a board-wide intervention,[6] with an expectation that such services had the potential to reduce risk from attendances to emergency care and high hospital occupancy.[16] The service received 7,000 monthly calls over its first 6 months.[6]

### Unit of analysis

The intervention was delivered to the health board, which is the unit of analysis. Board series were constructed by summing attendance and ≥4, ≥8, and ≥12 hour wait counts across constituent Type 1 sites and deriving percentages from those totals, so that each board series is attendance-weighted. Analysis at board level removes internal redistribution from the estimate; for example if FNC+ moved patients between EDs within the board, the board-level estimate provides a consistent estimate of whole-intervention effect on the number of attendances, while if this materially distributed demand to match available supply, it should be reflected by a reduction in long ED waits.

### Data and outcomes

Public Health Scotland publishes weekly, site-level ED data including attendances and the numbers of patients spending more than 4, 8 and 12 hours in an Emergency Department before admission, discharge or transfer.[12] Data were analysed from 1 January 2022 to one year after implementation (1 April 2025).

Attendance volume and the long-wait proportions were treated as co-primary outcomes; attendance volume being the principal outcome which FNCs have been stated to target, while the waits created from ED crowding are clinically-significant, with longer ED stays being associated with excess mortality.[1] Published thresholds are cumulative, every patient waiting over 12 hours also appears in the 8- and 4-hour counts.

Hospital occupancy and length of stay is associated with ED crowding,[17] and so changes in mean hospital length of stay may affect ED crowding, this is reported descriptively over the study period.

### Design and analysis

Interrupted time series analysis is widely used where randomisation is not feasible.[18] A controlled design augments this with contemporaneous control units, so that the counterfactual is anchored to shared changes affecting the whole system, for example seasonal pressures and national policy.[19]

The intervention effect was estimated as a level change at implementation, and slope change, expressed as that seen at one year following implementation. The difference between the treated board’s outcome and the mean of the control boards was regressed on a linear time trend and Fourier terms representing annual seasonality, with a binary intervention indicator.[20] Serial correlation was modelled with autoregressive moving average (ARMA 1,1) errors, and models were fitted by restricted maximum likelihood using generalised least squares.[21] Estimates are reported as percentage-point changes with 95% confidence intervals; attendance volume was modelled on the log scale and is reported as a proportional change.

### Controls

Control boards were selected using pre-intervention data only, so that selection could not be influenced by post-intervention outcomes. Candidates were ranked by the correlation of their pre-period residuals with those of the treated board, and the number retained chosen to minimise the Bayesian Information Criterion (BIC). All 13 other territorial health boards were eligible. The selected controls were NHS Grampian, NHS Fife, NHS Greater Glasgow & Clyde, NHS Tayside, NHS Lothian, NHS Ayrshire & Arran; derived from their similarity to the treated board on the 4-hour series and maintained for all comparisons.

### Sensitivity analyses

Because a complex intervention rarely achieves full effect at implementation,[18] transition sensitivity analyses were conducted: a washout excluding the first 4, 8 or 13 weeks, and a linear ramp reaching full effect over the same periods. A placebo falsification test was used, assigning treatment to six-months prior to the intervention.[18] Bayesian structural time series models were used as an independent estimator of effect. These used the same control pool, conservative local-level trends and weakly informative spike-and-slab priors.[22] Structural break assumptions were examined using an unsupervised Bayesian changepoint estimator,[23] and residuals assessed by Ljung–Box test and autocorrelation plots. Following the recommendation of Lopez Bernal et al.[19], a simple interrupted time series was also fitted.

### Precision and the interpretation of effect size

To determine the ability of the model to describe meaningful differences,[24] the design was simulated using pre-intervention data to give a minimum detectable effect at 80% power and alpha 0.05 of the value for the 4-hour wait outcome.[25] The minimum detectable effect, for change in the proportion waiting over 4 hours at the intervention board, was 2.4%. The clinically and operationally meaningful benefit was set at 5% for the wait thresholds and attendance volume; 95% confidence intervals lying entirely within ±5% were interpreted as excluding a clinically meaningful benefit.

### Analytic environment

As an evaluation of an already-implemented intervention using publicly available data, the study was defined as service evaluation and did not require ethical review.[26] Analyses used R version 4.5.0 (*R Core Team 2025*) with the nlme package.[21]

## Results

Between 1 January 2022 and 1 April 2025 there were 648,572 attendances across NHS Lanarkshire EDs, comprising 118 pre-intervention and 52 post-intervention weeks.

Summary characteristics are shown in Table 1.

**Table 1.** Attendances across NHS Lanarkshire EDs before and after implementation, by wait threshold. Percentages are derived from board totals and are therefore attendance-weighted across sites.

| Measure | Total | Pre | Post |
| --- | --- | --- | --- |
| Total Attendances | 648,572 | 446,780 | 201,792 |
| Total Attendances Per Week | 3,815 | 3,786 | 3,881 |
| Total Attendances Over 4 Hours | 277,747<br>(42.8%) | 187,174<br>(41.9%) | 90,573 (44.9%) |
| Total Attendances Over 8 Hours | 102,020<br>(15.7%) | 64,935 (14.5%) | 37,085 (18.4%) |
| Total Attendances Over 12 Hours | 42,965 (6.6%) | 25,413 (5.7%) | 17,552 (8.7%) |

ED attendances showed no detectable change (+1.20%, 95%CI −0.66 to +3.10%) relative to the counterfactual (Figure 1), with a confidence interval excluding a reduction as large as the meaningful difference of 5%. However, the slope change demonstrated that the effect increased across the one-year follow up; +3.95% (+0.36 to +7.67%) (Supplementary Table 1).

**Figure 1.**
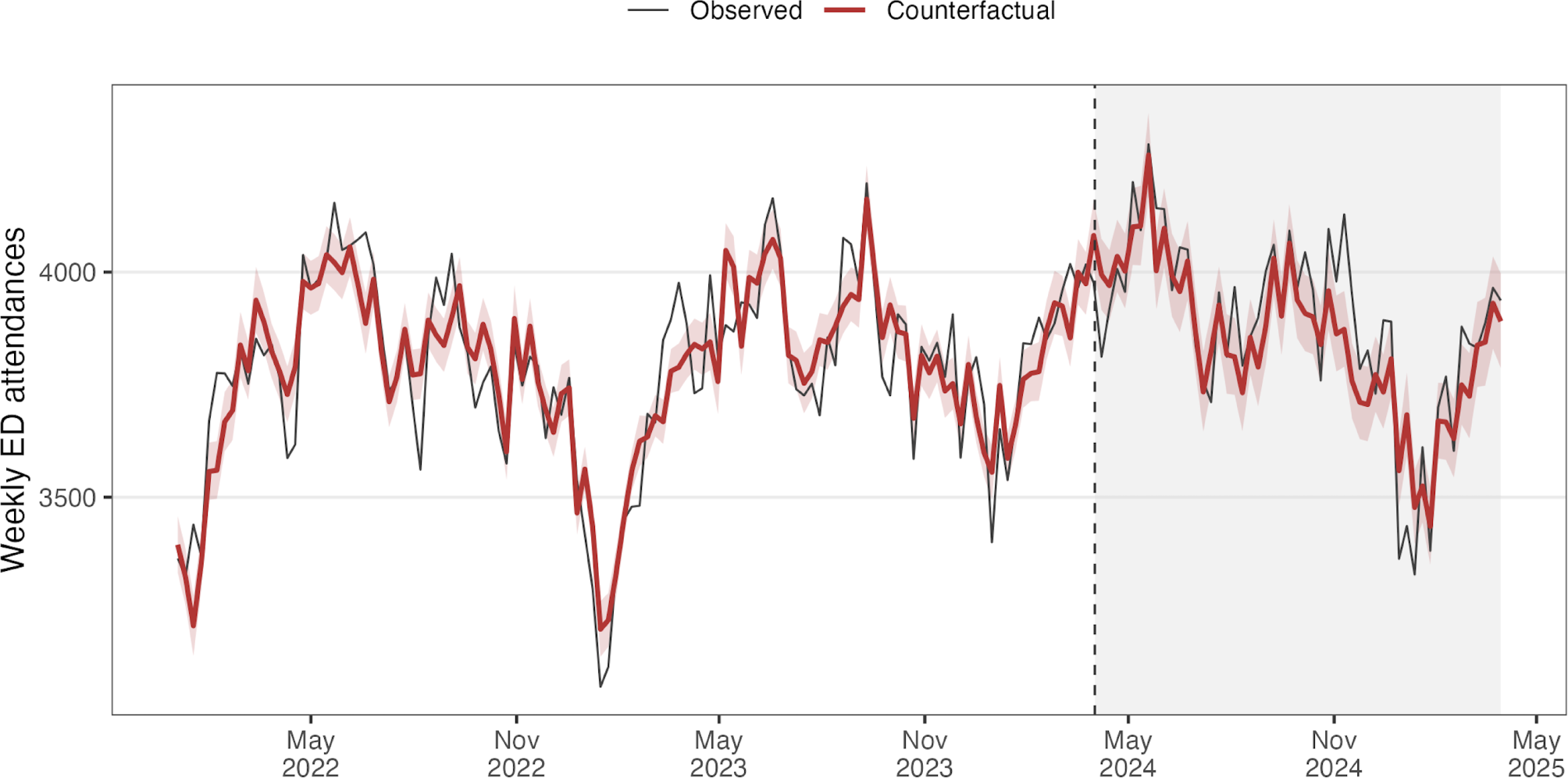
Weekly ED attendances, observed and counterfactual. Shaded band is the 95% confidence interval on the counterfactual; dashed line marks implementation.

The proportion of attendances waiting ≥4 hours showed no statistically significant change (+1.74%, 95%CI −0.71 to +4.20%) relative to the control-based counterfactual, albeit with a central estimate consistent with a deterioration in 4-hour performance. No significant differences were seen with the ≥8-hour series (+0.51%, 95% CI −1.86 to +2.88%), and the ≥12-hour series (−0.06%, 95%CI −1.71 to +1.59%). There were no significant changes to the slope for long waits. Effect estimates for all thresholds are given in Table 2 and visualised in Figure 2.

**Table 2.**
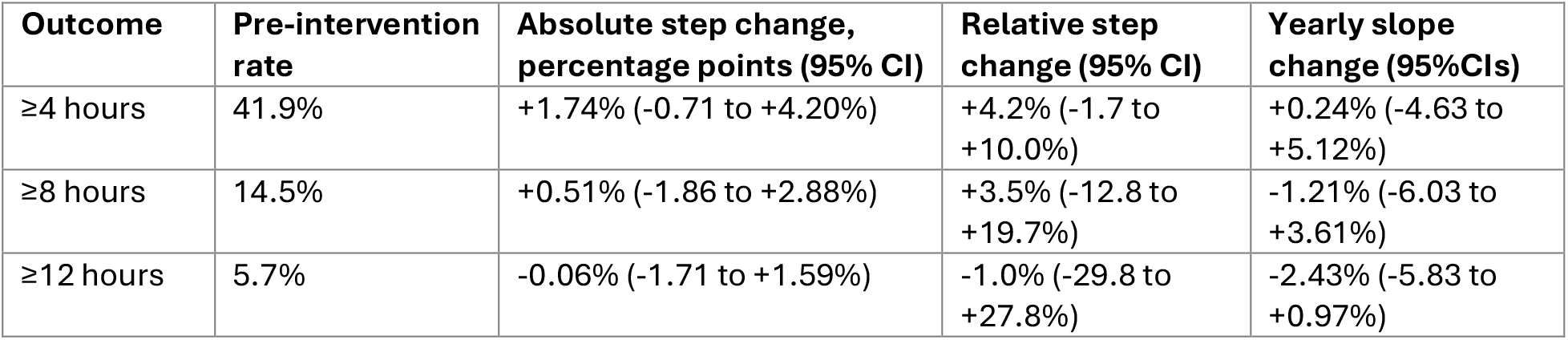
Change in the proportion of attendances breaching each threshold, expressed in absolute percentage points and relative to each series’ own pre-intervention rate.

| Outcome | Pre-intervention rate | Absolute step change, percentage points (95% CI) | Relative step change (95% CI) | Yearly slope change (95% CIs) |
| --- | --- | --- | --- | --- |
| ≥4 hours | 41.9% | +1.74% (-0.71 to +4.20%) | +4.2% (-1.7 to +10.0%) | +0.24% (-4.63 to +5.12%) |
| ≥8 hours | 14.5% | +0.51% (-1.86 to +2.88%) | +3.5% (-12.8 to +19.7%) | -1.21% (-6.03 to +3.61%) |
| ≥12 hours | 5.7% | -0.06% (-1.71 to +1.59%) | -1.0% (-29.8 to +27.8%) | -2.43% (-5.83 to +0.97%) |

**Figure 2.**
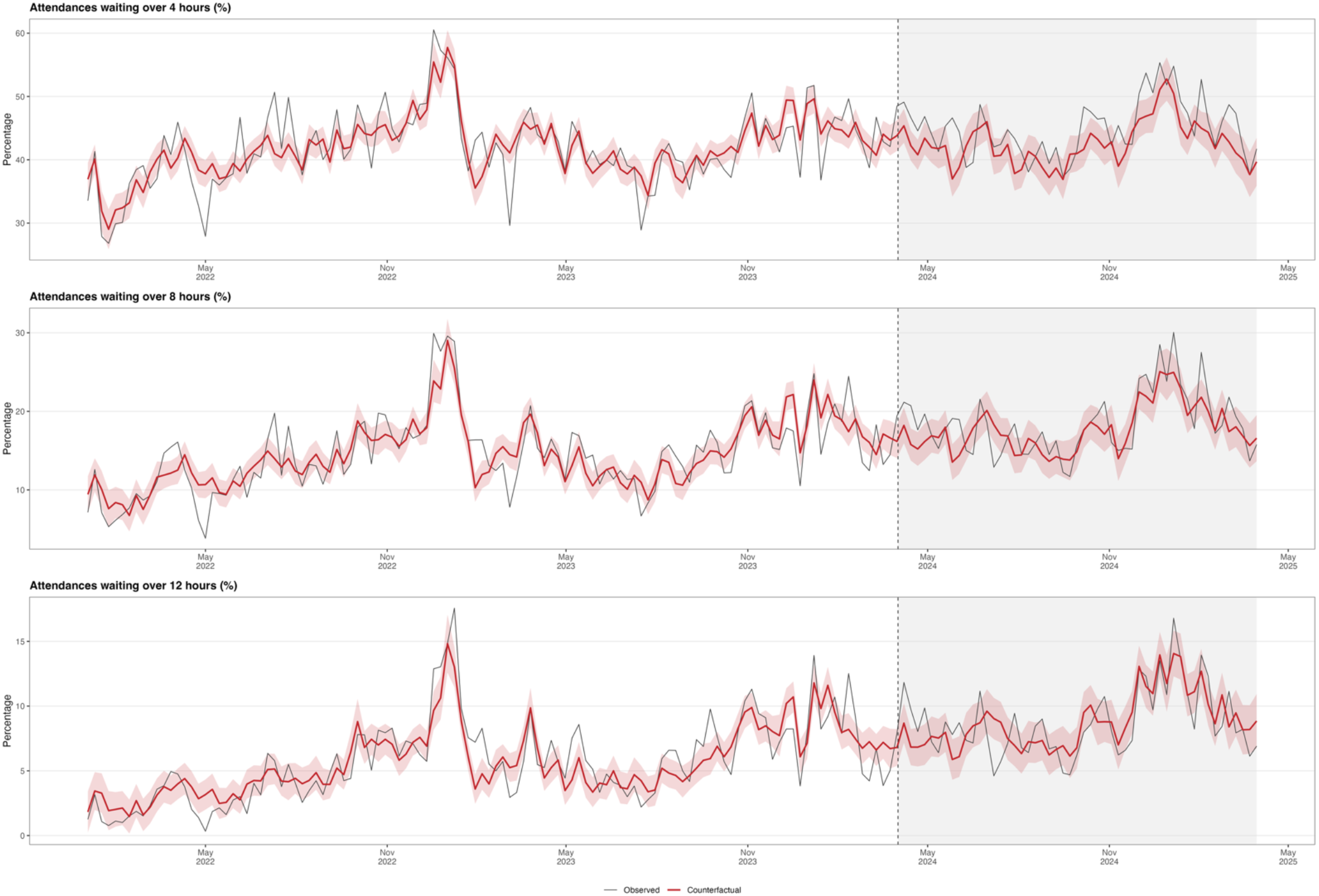
Proportion of ED attendances waiting ≥4 hours, ≥8 and ≥12 hours, observed and counterfactual. Shaded band is the 95% confidence interval on the counterfactual; dashed line marks implementation.

Mean hospital length of stay rose from 6.8 days in the first quarter of the study period to 8.5 days in the same quarter of 2025 (Supplementary Table 2 & Supplementary Figure 1).

There were no significant differences identified on simple single-board interrupted time series (Supplementary Table 3). Some washout and ramp specifications demonstrated significant increase in attendances following the intervention, for example those allowing a 13-week washout period following intervention demonstrated that the treated board had a 2.3% increase in attendances compared to the counterfactual (95%CIs +0.07 to +4.63%) (Supplementary Table 1).

Bayesian structural time series models demonstrated no significant change in total attendances, but a small increase in the percentage of 12-hour waits, with the average percentage increasing by 0.9% (95% Credible Interval 0.0 to 1.8%) (Supplementary Table 4 & Supplementary Figures 2A-D).

Structural break diagnostics indicated no concurrent structural breaks in the proportion of presentations waiting more than 4 hours in either the intervention or control sites. (Supplementary Figure 3). Ljung-Box test indicated no significant residual autocorrelation (p>0.05), and ACF/PACF plots demonstrated acceptable patterns of residual autocorrelation (Supplementary Figure 4). The placebo falsification tests demonstrated no significant differences irrespective of the washout or ramp specification for any outcome (p>0.05).

## Discussion

As-implemented, FNC+ was not associated with a change in ED attendances, nor with a change in the proportion of patients experiencing long waits. However, some sensitivity analyses and transition specifications demonstrated deterioration in delays and an increase in attendances. The primary central estimate did not exclude the potential that the 4-hour performance deteriorated in the year following the FNC’s introduction.

It is likely that the theory of supply-sensitive care supports some of the mechanism for these findings.[10] It is possible that the FNC+ operates as additional urgent care supply rather than as a filter on existing demand, and the degree to which that additional supply is high-value for patients and systems is unknown.

The finding of an increasing effect of attendances across the year of analysis aligns with those of sensitivity analyses allowing for a ramp or washout period; several of which also demonstrated increasing attendances following the intervention. These findings are contrary to the aims of the intervention, which sought to reduce self-presentation to the EDs. It is possible that remote assessment simultaneously diverted some patients who would otherwise have attended, and generated contacts from patients who may not otherwise have attended at all. Utilisation of services for which need is discretionary and capacity is the effective determinant tends to expand toward the capacity provided.[9,10] This finding is consistent with a comparable large-scale evaluation of the introduction of telephone-based medical advice in England, where such a service failed to reduce ED attendances or emergency admissions.[11] Further work is required to elucidate the exact patient-level mechanisms that can explain these findings.

Emergency Department crowding in systems of this kind is principally an exit problem, caused by the lack of inpatient capacity for patients who require it.[3,27] Interventions reducing arrivals are only likely to reduce crowding in systems constrained by demand.[7,8] Mean inpatient length of stay in the board rose across the study period, in contrast to other sites where interventions to reduce crowding have shown success.

### Strengths and limitations

The study benefits from use of nationally-reported, open data and methodology with demonstrated strengths in the evaluation of natural experiments.[28] The observed interval excludes a benefit as large as the meaningful difference of 5 percentage points, and cannot exclude a deterioration approaching the meaningful threshold, so this study supports the conclusion that FNC+ did not provide clinically-relevant reductions in either the volume of ED attendances or crowding.

The unit of analysis matches the intervention; evaluating a board-wide programme at a single site may confound its effect with redistribution between that site and its neighbours. The control selection, the transition and structural sensitivities address the main concerns to this methodological design.

The study has several limitations. Its use of board-wide, nationally-reported data aims to reflect changes to metrics reflecting the service’s aims, not individual patient journeys. It also does not reflect the fidelity of the intervention; ‘the degree to which an intervention or programme is delivered as intended’.[29] For example, the service may have been used by too few patients to impact the outcomes under review, or it may have been used by patients who would have attended regardless. Distinguishing these requires individual patient level data on healthcare use, the proportion directed to an ED, and understanding the counterfactual patient journey. These effects are not observable in routine published data.

The 4-, 8- and 12-hour thresholds are nested, so results for each should be interpreted as correlated rather than independent.

This is a quasi-experimental design and cannot exclude an unmeasured change coinciding with implementation that affected the treated board but not the controls. The FNC+ was implemented alongside other elements of the Redesign of Urgent Care programme, so the analysis estimates the effect of the package as delivered in this board rather than of FNC+ generally. Routine data carry no patient-level detail, so case mix and acuity are unobserved. One year of follow-up may be insufficient if the effect of FNC+ accumulates slowly or its initial implementation was limited by lack of staffing, low number of patient interaction or under-development of associated alternative care pathways.

## Conclusion

Board-wide implementation of an unscheduled care co-ordination hub, Flow Navigation Centre Plus, was not associated with an immediate significant change in emergency department long waits, and may be associated with increasing attendances as services are established. This evaluation does not exclude the potential for a deterioration in ED waits over 4 hours in the year following introduction of the FNC+. Such findings may be consistent with supply-sensitive care, where the introduction of a new route into urgent care consumes resource without meaningfully changing the requirements for care. There may be other benefits to individual patient journeys conferred by FNC+ that are not assessed here. Policymakers and healthcare managers should consider ensuring resource intended to reduce emergency care crowding is directed to its core causes; that of exit block and long waits for inpatient beds.

## Data Availability

All data is available hosted by Public Health Scotland

