## Supplement for "The implementation of an unscheduled care co-ordination hub (Flow Navigation Centre Plus), and emergency department attendances and delays: a controlled interrupted time series"

**Supplementary Appendix**

**Supplementary Table 1. Level change in percentage points (95%CI) for attendances and each waiting time threshold, by transition specification. 'Step' is the primary specification; 'washout' excludes the specified post-intervention weeks; 'ramp' phases the effect in linearly over the specified weeks.**

| Specification | Over 4 hours | Over 8 hours | Over 12 hours | Attendances |
|---|---|---|---|---|
| Step | 1.74 (-0.71 to 4.20) | 0.51 (-1.86 to 2.88) | -0.06 (-1.71 to 1.59) | 1.20 (-0.66 to 3.10) |
| Washout, 4 weeks | 1.54 (-1.00 to 4.08) | -0.12 (-2.57 to 2.34) | -0.83 (-2.59 to 0.94) | 1.86 (-0.07 to 3.82) |
| Washout, 8 weeks | 1.37 (-1.29 to 4.03) | -0.15 (-2.74 to 2.44) | -0.83 (-2.67 to 1.01) | 2.10 (-0.65 to 4.93) |
| Washout, 13 weeks | 1.54 (-1.27 to 4.34) | -0.51 (-3.33 to 2.31) | -1.24 (-3.25 to 0.77) | 2.33 (0.07 to 4.63) |
| Ramp, 4 weeks | 1.51 (-1.03 to 4.06) | -0.03 (-2.51 to 2.45) | -0.75 (-2.44 to 0.95) | 1.77 (-0.12 to 3.69) |
| Ramp, 8 weeks | 1.40 (-1.20 to 4.01) | -0.25 (-2.79 to 2.28) | -0.99 (-2.69 to 0.71) | 2.14 (0.24 to 4.07) |
| Ramp, 13 weeks | 1.20 (-1.47 to 3.87) | -0.57 (-3.15 to 2.02) | -1.25 (-2.96 to 0.46) | 2.33 (0.41 to 4.28) |

**Supplementary Table 2. Mean hospital length of stay in the intervention board across the study period.**

| Quarter | Mean length of stay (days) | Emergency stays |
|---|---|---|
| 2022 Q1 | 6.80 | 15,143 |
| 2022 Q2 | 6.91 | 15,662 |
| 2022 Q3 | 6.70 | 15,677 |
| 2022 Q4 | 7.03 | 15,833 |
| 2023 Q1 | 6.88 | 16,341 |
| 2023 Q2 | 6.58 | 15,790 |
| 2023 Q3 | 6.44 | 16,354 |
| 2023 Q4 | 7.17 | 15,516 |
| 2024 Q1 | 7.97 | 13,835 |
| 2024 Q2 | 7.70 | 13,611 |
| 2024 Q3 | 8.12 | 13,232 |
| 2024 Q4 | 8.20 | 13,335 |
| 2025 Q1 | 8.54 | 12,998 |
| 2025 Q2 | 7.86 | 13,590 |

**Supplementary Figure 1. Mean inpatient length of stay for the intervention board across the study period. The quarter in which the intervention commenced is noted in the grey band.**

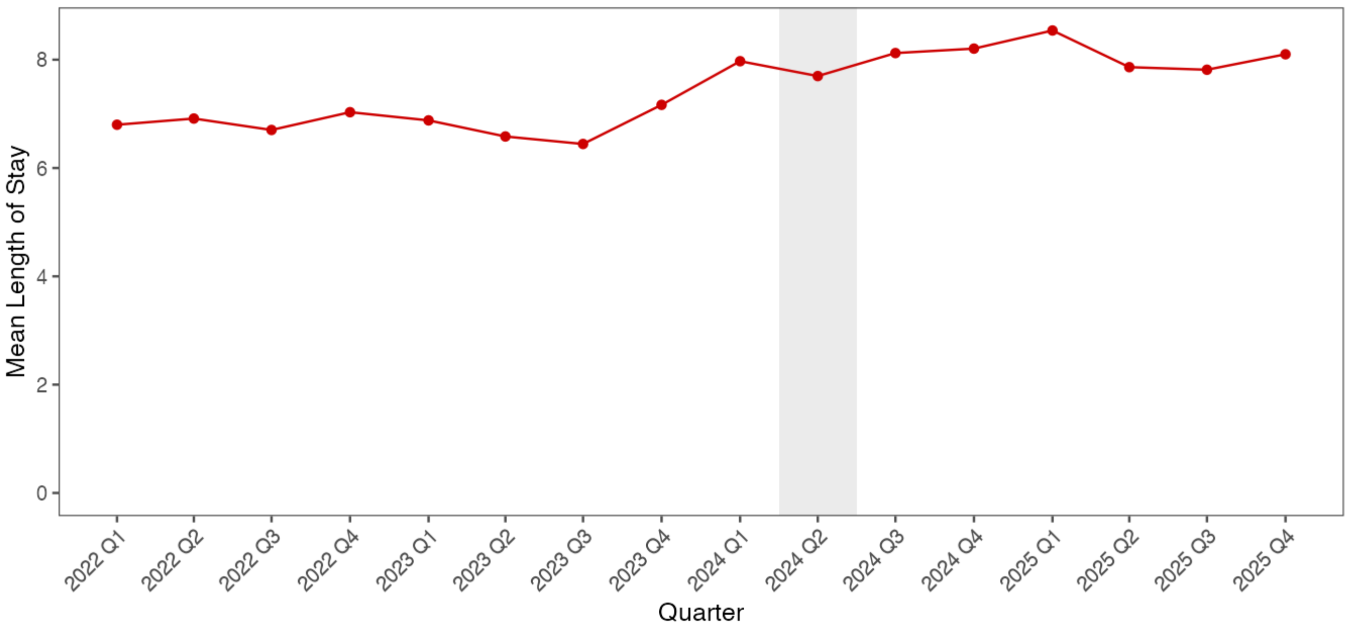

**Supplementary Table 3. Results of simple interrupted time series analysis on the effect of the intervention to existing trends within the intervention board.**

| Outcome | Step Change | Slope Change (per week) |
|---|---|---|
| **Wait ≥4 hours** | 1.12% (95%CI -6.13 to 8.37%) | -0.07% (95%CI -0.33 to 0.18%) |
| **Wait ≥8 hours** | 1.74% (95%CI -3.70 to 7.18%) | -0.08% (95%CI -0.26 to 0.11%) |
| **Wait ≥12 hours** | 1.91% (95%CI -1.12 to 4.93%) | -0.07% (95%CI -0.17 to 0.02%) |
| **Attendances** | -2.94% (95%CI -8.41 to 2.53%) | 0.02% (95%CI -0.17 to 0.20%) |

**Supplementary Table 4. Bayesian structural time-series posterior estimates for the effect of the intervention using the control pool used in the primary analysis, with posterior tail-area probabilities expressed as p-values.**

| Outcome | Unit | Average Change (95% Credible Interval) | Posterior tail-area probabilities |
|---|---|---|---|
| **Total Attendances** | Weekly attendances | -5.5 (-82.3 to 42.4) | 0.441 |
| **Wait ≥4 hours** | Percentage Point | +0.9% (-0.7 to 2.7%) | 0.155 |
| **Wait ≥8 hours** | | +1.1% (-0.2 to 2.4%) | 0.061 |
| **Wait ≥12 hours** | | +0.9% (0.0 to 1.8%) | 0.029 |

**Supplementary Figures 2A-D. Plots of Bayesian structural time series analysis of the effect of the intervention on the number of patients waiting ≥4 (A), ≥8 (B), and ≥12 hours (C), and number of attendances (D), following the intervention.**

**2A**

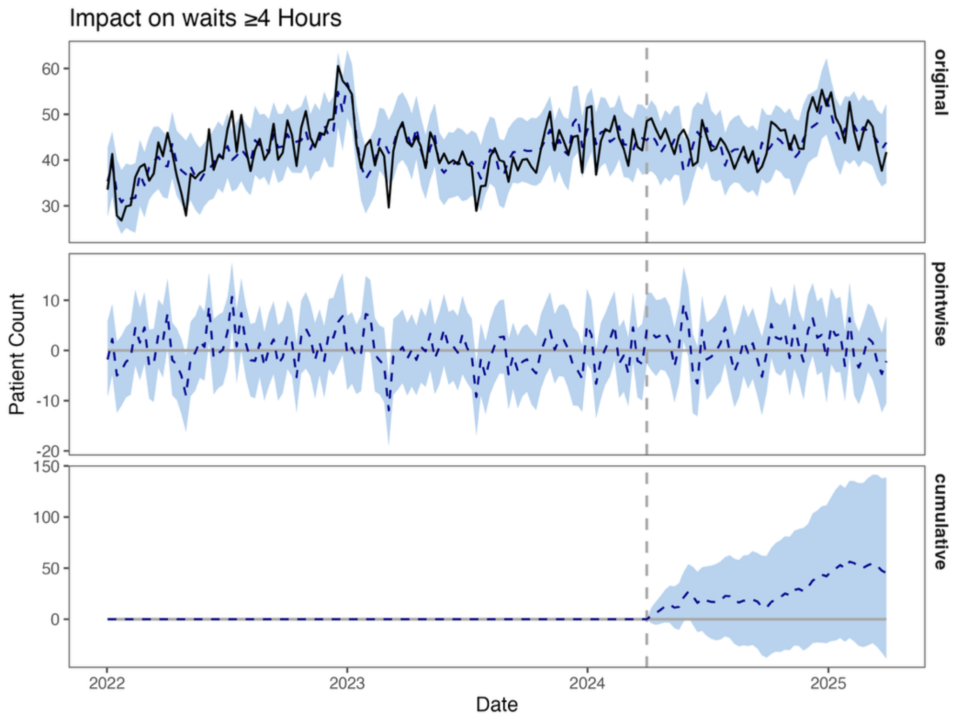


**2B**

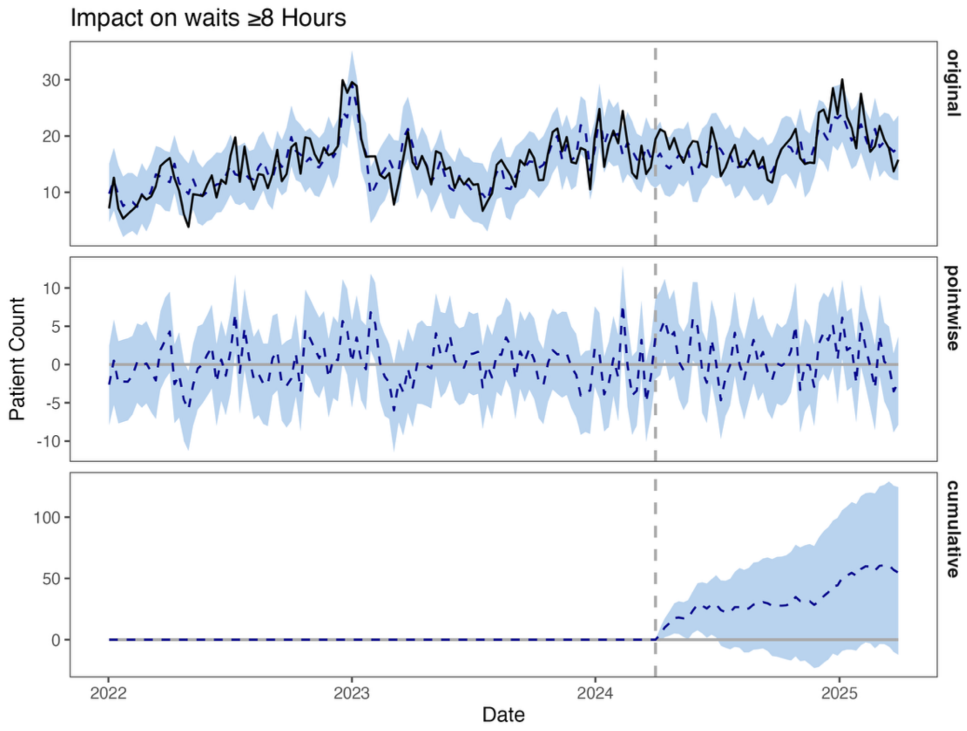

## 2C

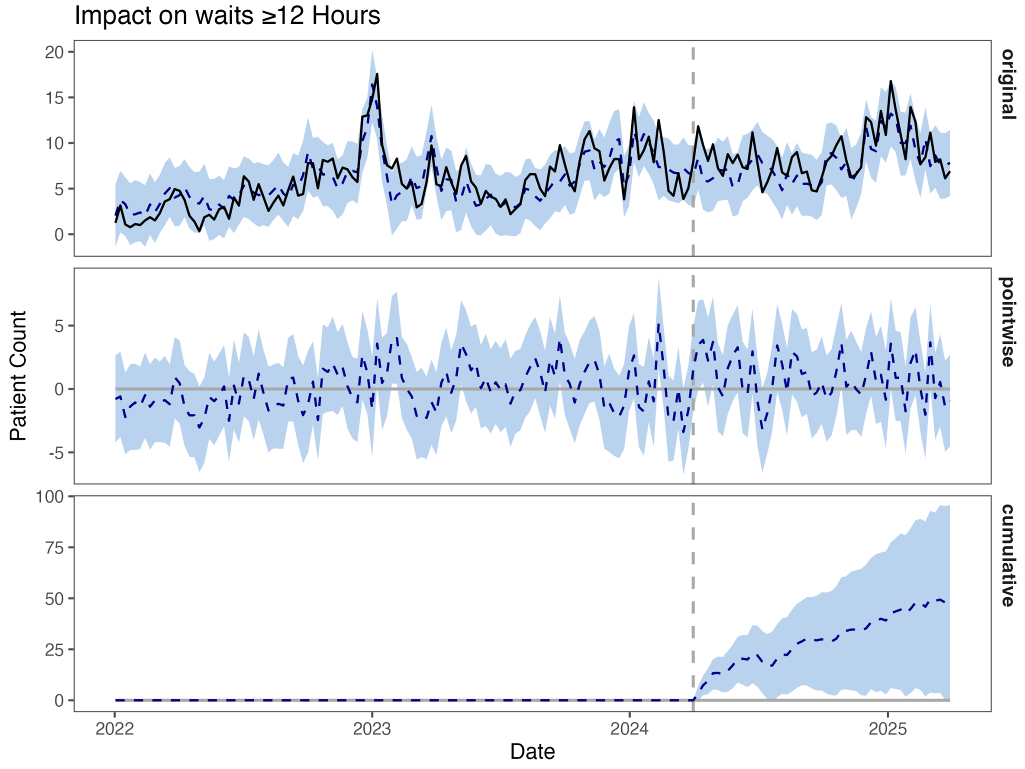
Impact on waits ≥12 Hours
original
pointwise
cumulative
Patient Count
Date
2022
2023
2024
2025
20
15
10
5
0
-5
100
75
50
25

## 2D

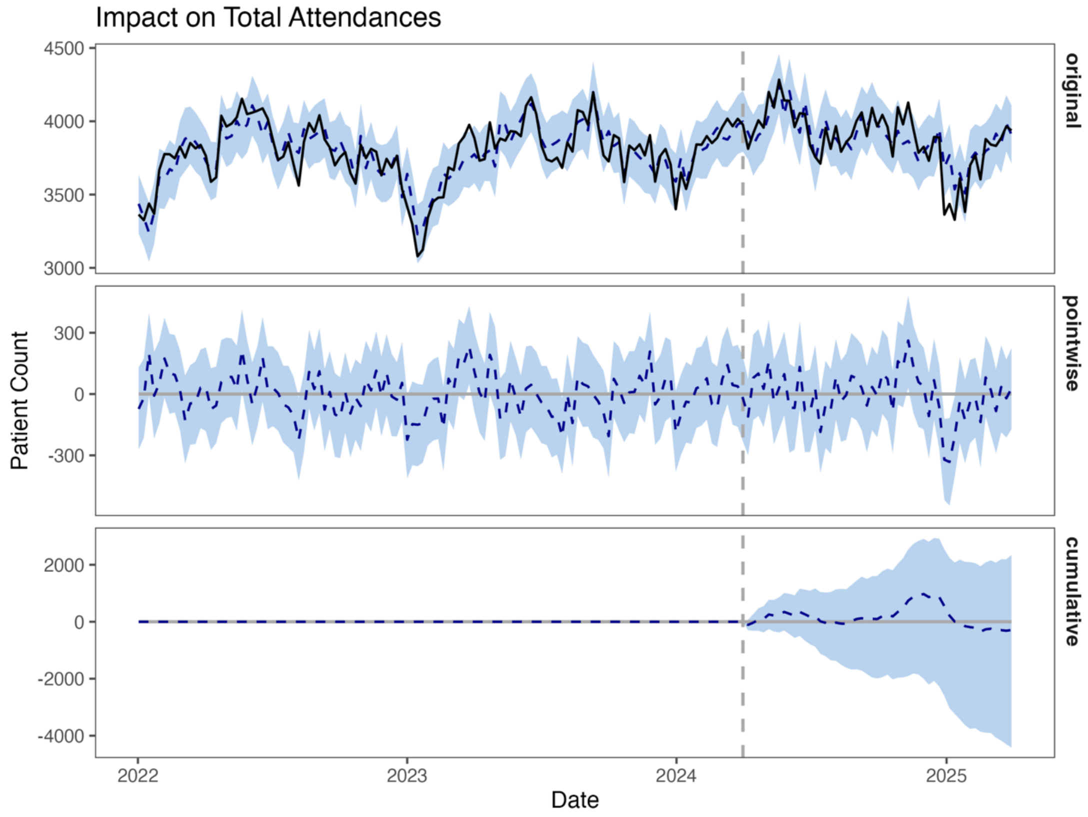
Impact on Total Attendances
original
pointwise
cumulative
Patient Count
Date
4500
4000
3500
3000
300
0
-300
2000
-2000
-4000
2022
2023
2024
2025

**Supplementary Figure 3. Bayesian structural break diagnostics.**

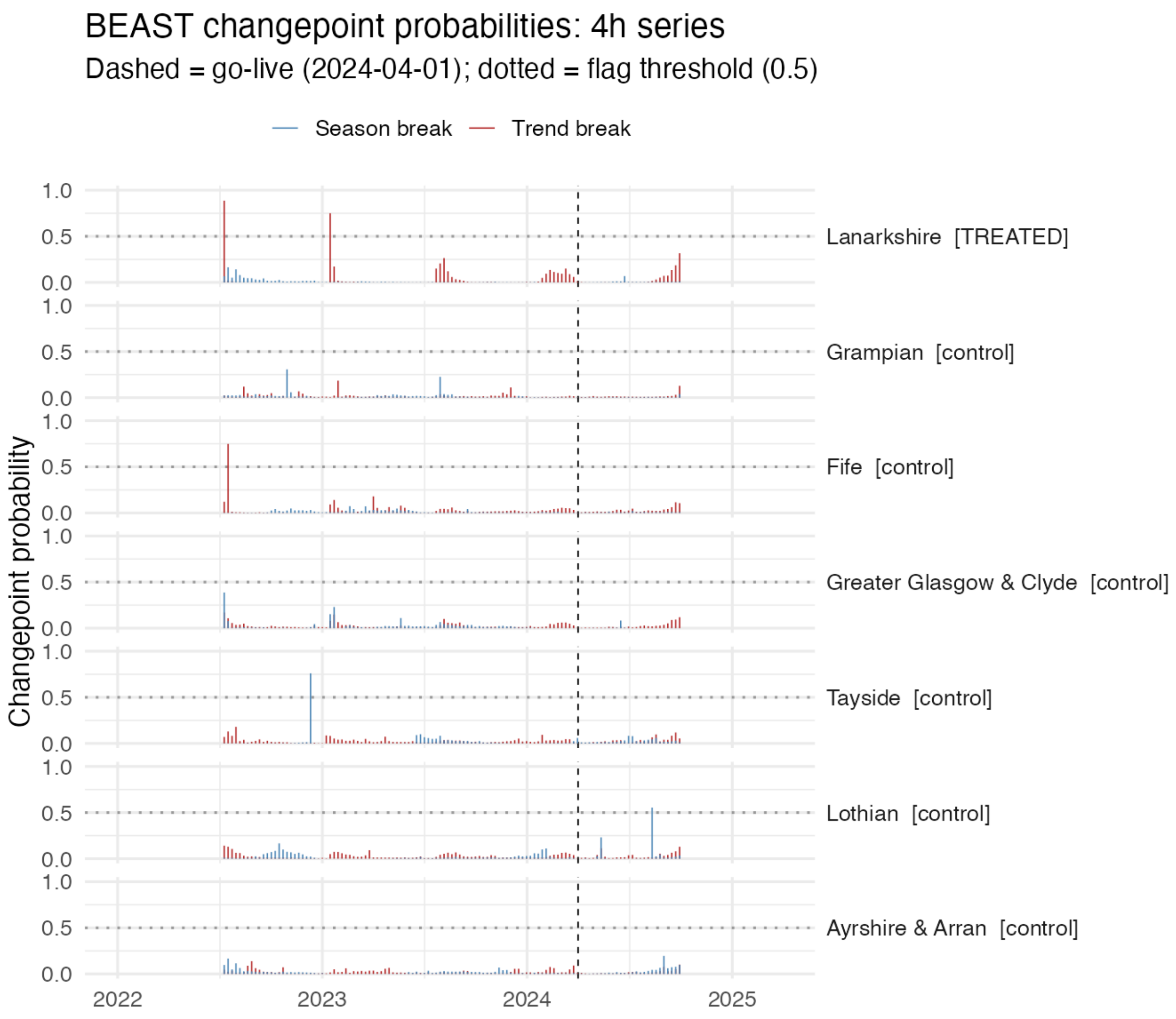

**Supplementary Figure 4. Autocorrelation function and partial autocorrelation function plots for attendances and each waiting time threshold.**

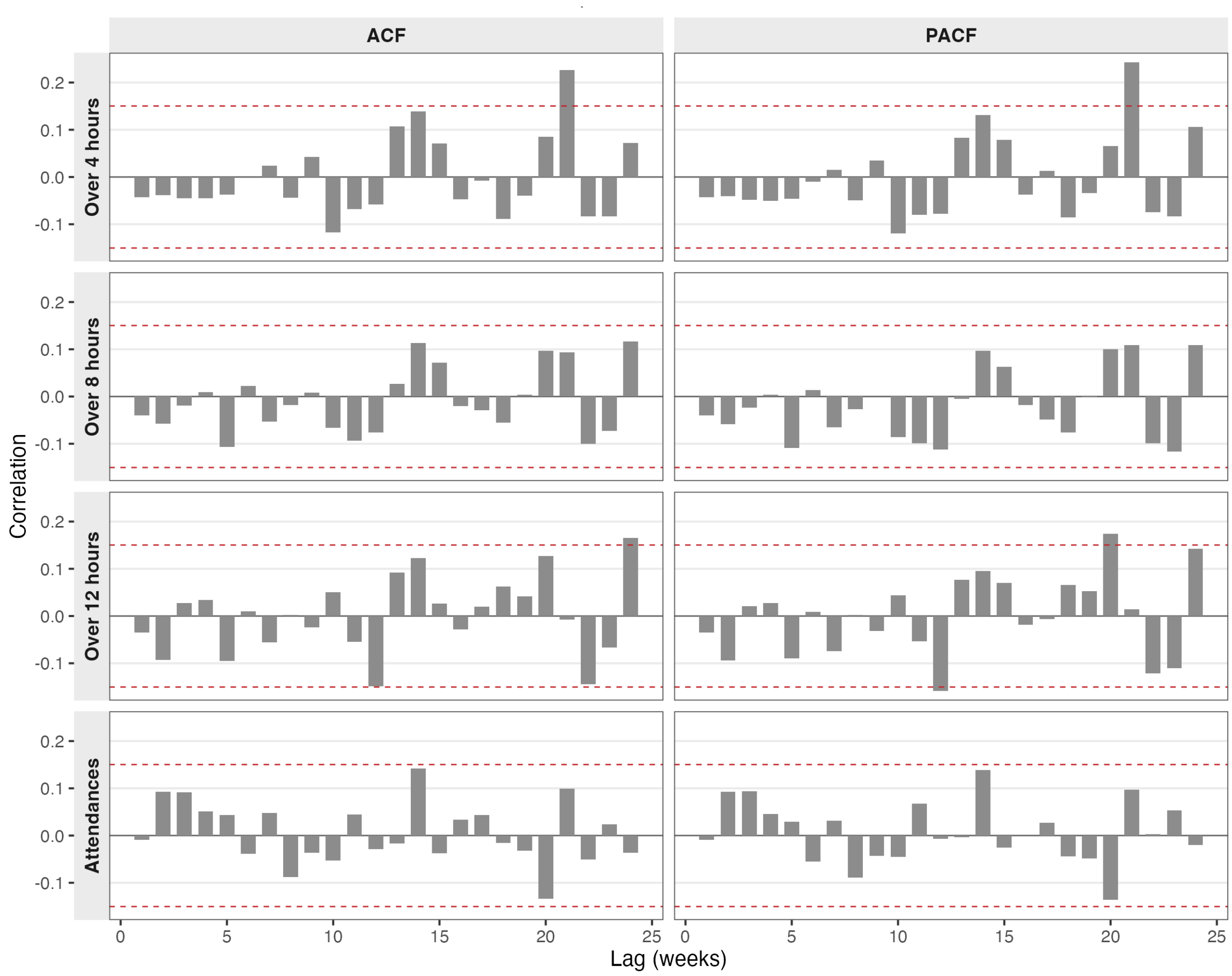
